# Post COVID-19 condition after Wildtype, Delta, and Omicron variant SARS-CoV-2 infection and vaccination: pooled analysis of two population-based cohorts

**DOI:** 10.1101/2022.09.25.22280333

**Authors:** Tala Ballouz, Dominik Menges, Marco Kaufmann, Rebecca Amati, Anja Frei, Viktor von Wyl, Jan S. Fehr, Emiliano Albanese, Milo A. Puhan

## Abstract

**Background:** Post COVID-19 condition (PCC) is an important complication of SARS-CoV-2 infection, affecting millions worldwide. Further evidence is needed on the risk of PCC after vaccination and infection with newer variants. This study aimed to evaluate the prevalence and severity of PCC across different variants and vaccination histories.

**Methods:** We used pooled data from 1350 SARS-CoV-2-infected individuals from two representative population-based cohorts in Switzerland, diagnosed between Aug 5, 2020, and Feb 25, 2022. We descriptively analysed the prevalence and severity of PCC, defined as the presence and frequency of PCC-related symptoms six months after infection, among vaccinated and non-vaccinated individuals infected with Wildtype, Delta, and Omicron SARS-CoV-2. We used multivariable logistic regression models to assess the association and estimate the risk reduction of PCC after infection with newer variants and prior vaccination. We further assessed associations with the severity of PCC using multinomial logistic regression. To identify groups of individuals with similar symptom patterns and evaluate differences in the presentation of PCC across variants, we performed exploratory hierarchical cluster analyses.

**Findings:** We found strong evidence that vaccinated individuals infected with Omicron had a reduced risk of developing PCC compared to non-vaccinated Wildtype-infected individuals (odds ratio 0.42, 95% confidence interval 0.24–0.68). The risk among non-vaccinated individuals was similar after infection with Delta or Omicron compared to Wildtype SARS-CoV-2. We found no differences in PCC prevalence with respect to the number of received vaccine doses or timing of last vaccination. The prevalence of PCC-related symptoms among vaccinated, Omicron-infected individuals was lower across severity levels. In cluster analyses, we identified four clusters of diverse systemic, neurocognitive, cardiorespiratory, and musculoskeletal symptoms, with similar patterns across variants.

**Interpretation:** The risk of PCC appears to be lowered with infection by the Omicron variant and after prior vaccination. This evidence is crucial to guide future public health measures and vaccination strategies.

**Funding:** Swiss School of Public Health (SSPH+), University of Zurich Foundation, Cantonal Department of Health Zurich, Swiss Federal Office of Public Health

**Study registrations:** **<u>ISRCTN14990068</u>, <u>ISRCTN18181860</u>**

**Research in context:** *Evidence before this study:* We searched MEDLINE, EMBASE, and medRxiv for primary studies assessing the prevalence and symptoms associated with post COVID-19 condition (PCC) after infection with different SARS-CoV-2 variants and among infected individuals with and without prior vaccination. We used a specific search strategy limited to the timeframe between Jan 01, 2020, and Aug 29, 2022, without language restriction (reported in Supplementary Table S1). We further searched identified systematic reviews for additional references. We screened 221 unique records and identified four studies investigating the association of Delta or Omicron variant infections and 11 studies investigating the association of prior vaccination with PCC. Current evidence is uncertain whether infection with emerging variants may be associated with a reduction of the risk of developing PCC. Two studies found a decreased risk of PCC with Omicron compared to Delta infection, or to individuals infected during any prior wave. One study found a lower risk of PCC with Alpha compared to Wildtype SARS-CoV-2, but an increased risk among those infected with the Delta or Omicron variant. One study primarily examined symptom clusters across different waves. All identified studies defined PCC as symptoms occurring at ≥4 weeks or ≥12 weeks after infection, and were conducted among hospitalised patients, healthcare workers, or users of the United Kingdom ZOE symptom app. Evidence regarding the preventive effects of vaccination on PCC was of higher certainty, with eight out of 11 studies reporting a substantially reduced PCC incidence with mRNA- and adenovirus vector-based vaccines. The magnitude of the effect in these studies varied, with estimated adjusted odds ratios ranging from 0.22 to 0.85. Nonetheless, three studies found no difference between vaccinated and non-vaccinated infected individuals, including two of three studies evaluating PCC at six months after infection. The third study with a six-month horizon found a higher odds ratio than any other study reporting a reduction at ≥4 weeks or ≥12 weeks. Study populations and designs varied strongly, and only one study evaluated the independent effects of SARS-CoV-2 variants and vaccination.

*Added value of this study:* This study investigates the association of PCC with infection with Delta and Omicron variants and prior vaccination compared to Wildtype SARS-CoV-2 infection among unvaccinated individuals. We found that infection with the Omicron variant and prior vaccination were associated with a lower risk of developing PCC six months after infection. Compared to previous work, this study is the first to evaluate PCC with a longer-term follow-up, while simultaneously evaluating the risk reduction by Delta and Omicron variants and prior vaccination on PCC. By relying on prospectively collected data from two representative population-based cohorts, we were able to provide an in-depth analysis of the longer-term risk reduction through prior vaccination and novel variants, the severity of PCC-related symptoms, and symptom clusters across pandemic waves between 2020 and early 2022.

*Implications of all the available evidence:* In conjunction with existing evidence, our study suggests that infection with the Omicron variant and prior vaccination are likely to substantially reduce the risk of developing PCC compared to infection with Wildtype SARS-CoV-2 without prior vaccination. We demonstrate that this risk reduction persists up to six months after infection, and that PCC-related symptoms are reduced to a similar extent across different levels of severity. However, the risk of having mild to even potentially severe PCC six months after infection is not eliminated. Hence, vaccinations will likely continue to be an important mainstay in the management of the further course of the pandemic. The prevention of further infection and PCC may still provide important benefits for public health, even if SARS-CoV-2 further evolves to cause milder infections and becomes endemic. Therefore, information from this study will be crucial to guide vaccination strategies and decisions on timing and enforcement of public health measures worldwide.

## Introduction

Post COVID-19 condition (PCC) is an important complication of SARS-CoV-2 infection, posing a substantial burden on health care systems worldwide.^1,2^ Population-based studies have estimated that about 20–30% of non-vaccinated individuals with a confirmed Wildtype SARS-CoV-2 infection develop PCC,^3^ defined as symptoms persisting beyond three months after infection and not explained by an alternative diagnosis.^1^ As of writing, half a billion SARS-CoV-2 infections have been diagnosed worldwide,^4^ leading to more than 144 millions of people estimated to have been affected by PCC in 2020 and 2021.^2^ While most individuals with PCC are mildly affected and recover within months after infection,^2,5^ some suffer from protracted symptoms with a substantial impact on daily activities, social and professional life.^3^ With the global implementation of vaccination campaigns and the emergence of novel SARS-CoV-2 variants of concern, future management of the pandemic will thus likely depend on the incidence of PCC in vaccinated individuals infected with novel variants.

Current evidence suggests that the risk to develop PCC is relevantly reduced by vaccination with mRNA-based or adenovirus vector-based vaccines.^6^ Several studies have been conducted using different study designs and populations, with the majority finding that the risk of PCC is approximately halved in vaccinated compared to non-vaccinated individuals.^7–17^ While most studies evaluated symptoms persisting for more than four weeks^8,9,17^ or more than 12 weeks^10,12,13,15,16^ after SARS-CoV-2 infection, only three assessed symptoms beyond six months and identified a lower^7^ or no^11,14^ risk reduction with vaccination.

Meanwhile, the evidence regarding infection with novel SARS-CoV-2 variants remains limited and inconsistent. While there is some evidence that infections with the Omicron variant reduced the risk of developing PCC compared to the Delta^18^ or any previous variant^19^, evidence from another study demonstrated a reduced risk with the Alpha variant, but not with the Delta or Omicron variant compared to Wildtype SARS-CoV-2.^9^ However, all three studies drew from specific samples not representative of the general population, and none of them has evaluated PCC for more than 12 weeks after infection. Therefore, more knowledge regarding the expected risk reduction through infection with newer variants and the preventive effects of vaccination on PCC in the longer-term is urgently needed.

This study aimed to evaluate the prevalence and severity of PCC in individuals infected by Wildtype, Delta, and Omicron SARS-CoV-2 with and without prior vaccination. Specific objectives were to assess the difference in risk of developing PCC with emerging variants and vaccination, evaluate changes in symptom severity, and identify prevalent symptom clusters and their evolution across pandemic waves. Thereby, we aim to inform public health policies and vaccination strategies worldwide for the future management of the pandemic.

## Methods

### Study design and participants

This study is based on a pooled analysis using data from two population-based cohorts in Switzerland (Supplementary Table S2). The Zurich SARS-CoV-2 Cohort is a prospective longitudinal cohort of 1106 SARS-CoV-2-infected individuals, recruited shortly after infection based on an age-stratified random sample of all diagnosed cases between Aug 6, 2020, and Jan 19, 2021, through the Department of Health of the canton of Zurich, Switzerland.^5,20^ The study was preregistered (https://doi.org/10.1186/ISRCTN14990068) and approved by the ethics committee of the canton of Zurich (BASEC 2020-01739). The Corona Immunitas seroprevalence study is a prospective longitudinal cohort of an age-stratified random population sample derived through the Swiss Federal Statistical Office.^21,22^ For this study, we leveraged data from the fifth phase of Corona Immunitas and associated follow-up from the cantons of Zurich (largest canton in German-speaking north-eastern Switzerland with around 18% of the Swiss population), and Ticino (Italian-speaking part with about 4% of Swiss population), Switzerland, including 1844 participants, for which baseline assessments took place between Mar 1–31, 2022.^23^ The study was registered (https://doi.org/10.1186/ISRCTN18181860) and approved by the ethics committees of the cantons of Zurich (BASEC 2020-01247) and Ticino (BASEC 2020-01514). All participants provided informed consent prior to participation.

### Data collection and pooling

In both cohorts, we collected data electronically using online survey questionnaires. The questionnaires of the two cohorts were closely aligned and used the same wording for critical questions related to sociodemographic characteristics, current symptoms, and current health status. The presence of PCC-related symptoms was elicited using a list of 23 symptoms frequently reported in the literature (Supplementary Table S3). We collected and managed all data through the Research Electronic Data Capture (REDCap) platform.^24,25^

In the Zurich SARS-CoV-2 Cohort, participants completed a baseline questionnaire shortly after infection including questions on sociodemographic characteristics, medical comorbidities, and initial SARS-CoV-2 infection. They were followed-up repeatedly over time to elicit data on symptom trajectories, medical complications, current health status, and new infections and vaccinations.^5,26^ From this cohort, we included all participants who had completed follow-up at six months after SARS-CoV-2 infection (a range of five to seven months after infection was allowed; Supplementary Figure S1).

In Corona Immunitas, participants equally completed a baseline questionnaire including questions on sociodemographic characteristics, medical comorbidities, received vaccinations, past SARS-CoV-2 infections, current PCC-related symptoms, and current health status. They were followed-up at two and four months after baseline to elicit data on PCC-related symptoms, current health status, new infections, and vaccinations.^5,21,23^ For this study, we included all participants who, either at baseline or during follow-up, reported having last been infected by SARS-CoV-2 during the timeframes of predominance of the Delta and Omicron variants (see definition below), and who completed the baseline or either of the follow-up assessments approximately six months after the most recent reported infection (five to seven months allowed; Supplementary Figure S1).

### Outcome definition

We defined the primary outcome of PCC as symptoms present within the last seven days at six months after the most recent diagnosed SARS-CoV-2 infection, which were reported by participants to be related to COVID-19. Based on prior research, prevalence of PCC-related symptoms approximately corresponds to the absolute risk difference of symptoms among infected compared to uninfected individuals from the general population.^5^ Secondary outcomes were individual symptoms reported to be related to COVID-19 and severity of PCC. We assessed severity using current symptom count at follow-up, stratified into groups with one to two, three to five, and six or more symptoms. In sensitivity analyses, we categorised patients based on the EuroQoL visual analogue scale (EQ-VAS) and previously applied cut-offs^5,27,28^ as having mild (EQ-VAS >70), moderate (EQ-VAS 51–70), or severe (EQ-VAS ≤50) PCC.

In the absence of viral samples for genetic analysis of SARS-CoV-2 variants, we determined the most likely infecting variant based on reported infection dates. We categorised all Zurich SARS-CoV-2 Cohort participants (diagnosed between Aug 5, 2020, and Jan 19, 2021) as infected by Wildtype SARS-CoV-2. In accordance with viral predominance (≥80% of diagnosed infections) in Switzerland, we determined infections between Jul 7, 2021, and Dec 31, 2021, as most likely Delta infections, and infections from Jan 1, 2022, as most likely Omicron infections.^29^ We used the date of the first positive SARS-CoV-2 test as infection date, while instances of a positive test more than 60 days before were considered a separate prior infection in analyses.

### Statistical analysis

We descriptively analysed population characteristics and the prevalence of PCC, overall and stratified by SARS-CoV-2 variants, prior vaccination, and symptom severity. We calculated 95% Wilson confidence intervals (CIs) for reported proportions. We further used multivariable logistic regression models to evaluate the association of infection during different variant timeframes and vaccination with the presence of PCC. In the primary analysis model, we combined variant and vaccination status information to evaluate the joint association within vaccinated or non-vaccinated, Delta or Omicron SARS-CoV-2-infected individuals compared to Wildtype infection among non-vaccinated individuals, while adjusting for prior infection. Model selection was based on prior knowledge (a priori variables were age, sex, presence of comorbidities (any of hypertension, diabetes, cardiovascular disease, chronic respiratory disease, malignancy, or immune suppression), and initial hospitalization due to COVID-19) and the Akaike and Bayesian Information Criteria (AIC/BIC; no further variables were added). In sensitivity analyses and to ensure comparability with other studies,^9,19^ we used an alternative model specification including SARS-CoV-2 variant, vaccination status, and prior infection as separate variables. We calculated odds ratios (ORs) and associated 95% CIs for all variables of interest. Additionally, we calculated the absolute risk reduction (i.e., adjusted risk differences) with 95% CI based on a logistic regression model using an identity link.^30^ Furthermore, we conducted multivariable multinomial logistic regression analyses to evaluate the association of different strains with the severity of PCC. We performed sensitivity analyses using alternative definitions for the severity of PCC.

In exploratory analyses, we assessed the presence of specific clusters of PCC-related symptoms at six months using multiple correspondence combined with hierarchical cluster analyses.^31^ Sixteen long-term symptoms reported by at least five percent of participants with PCC were included, on which a multiple correspondence analysis was performed. We retained dimensions that explained 90% of the variance and performed an agglomerative hierarchal clustering on the selected dimensions using Ward minimum-variance linkage methods.^31,32^ We based the selection of the number of clusters on findings from other studies^33–35^ and by maximizing the relative loss of inertia. We selected four clusters for the main analysis, but present findings from sensitivity analyses assuming five and six clusters for future comparison. We descriptively evaluated PCC-related symptoms and participant characteristics across clusters to identify factors potentially associated with membership in each of the clusters.

All analyses were performed in R (version 4.0.3).

## Results

We included data from 1045 Zurich SARS-CoV-2 Cohort participants and 305 Corona Immunitas participants reporting a SARS-CoV-2 infection with six months of follow-up (Table 1, Supplementary Figure S1, Supplementary Tables S4–5). Median follow-up was 183 days (interquartile range [IQR] 182– 186 days) across all participants. Zurich SARS-CoV-2 Cohort participants were slightly older on average, with a median age of 51 years (IQR 35–66) compared to 43 years (30–54) among Corona Immunitas participants. The proportion of female participants was 50.7% and 58.7%, respectively. Participants more frequently reported the presence of at least one medical comorbidity in the Zurich SARS-CoV-2 Cohort (29.5%) compared to Corona Immunitas (14.1%). 232 (77.1%) Corona Immunitas participants reported to have received at least one COVID-19 vaccine prior to infection, among which 173 (74.6%) had received one or two doses, and 59 (25.4%) had received three doses. Almost all (n=231, 99.6%) had received a mRNA-based vaccine (i.e., BNT162b2 or mRNA-1273) while one participant had received a vector-based vaccine (i.e., JNJ-78436735). A previous SARS-CoV-2 infection was reported by 36 (11.8%) of Corona Immunitas participants.

**Table 1:** Participant characteristics of Wildtype, Delta, or Omicron SARS-CoV-2-infected individuals from the Zurich SARS-CoV-2 Cohort and the Corona Immunitas Phase 5 seroprevalence studies. BMI = body mass index, IQR = interquartile range. * Percentages among those that have received at least one vaccine dose prior to infection.

|  | **Zurich SARS-CoV-2 Cohort** | **Corona Immunitas** | **Overall** |
| --- | --- | --- | --- |
|  | **(N=1045)** | **(N=305)** | **(N=1350)** |
| **Timeframe of diagnosis** | Aug 5, 2020 – Jan 19, 2021 | Jul 15, 2021 – Feb 25, 2022 | Aug 5, 2020 – Feb 25, 2022 |
| **Median follow-up (IQR; days)** | 183.5 (182–186) | 181 (164–196) | 183 (182–186) |
| **Age, median (IQR)** | 51 (35–66) | 43 (30–54) | 48 (34–63) |
| **Female sex** | 530 (50.7%) | 179 (58.7%) | 709 (52.5%) |
| **Presence of chronic comorbidity** | 308 (29.5%) | 43 (14.1%) | 351 (26.0%) |
| **Smoking status** |  |  |  |
| Non-smoker | 625 (60.1%) | 201 (65.9%) | 826 (61.4%) |
| Ex-smoker | 282 (27.1%) | 62 (20.3%) | 344 (25.6%) |
| Smoker | 133 (12.8%) | 42 (13.8%) | 175 (13.0%) |
| *Missing* | 5 | 0 | 5 |
| **Body mass index, median (IQR; kg/m<sup>2</sup>)** | 24.2 (21.9–26.6) | 23.3 (21.5–26.0) | 24.0 (21.7–26.5) |
| **Highest education** |  |  |  |
| None or mandatory school | 41 (3.9%) | 18 (5.9%) | 59 (4.4%) |
| Vocational training or specialised baccalaureate | 438 (42.2%) | 154 (50.8%) | 592 (44.1%) |
| Higher technical school or college | 276 (26.6%) | 36 (11.9%) | 312 (23.2%) |
| University | 284 (27.3%) | 95 (31.4%) | 379 (28.2%) |
| *Missing* | 6 | 2 | 8 |
| **Employment status** |  |  |  |
| Employed | 668 (64.2%) | 57 (18.8%) | 725 (53.9%) |
| Retired | 256 (24.6%) | 181 (59.5%) | 437 (32.5%) |
| Student | 50 (4.8%) | 32 (10.5%) | 82 (6.1%) |
| Unemployed or other | 67 (6.4%) | 34 (11.2%) | 101 (7.5%) |
| *Missing* | 4 | 1 | 5 |
| **Hospitalised due to COVID-19** | 44 (4.2%) | 2 (0.7%) | 46 (3.4%) |
| **SARS-CoV-2 variant** |  |  |  |
| Wildtype | 1045 (100%) | 0 (0.0%) | 1045 (77.4%) |
| Delta | 0 (0.0%) | 99 (32.5%) | 99 (7.3%) |
| Omicron | 0 (0.0%) | 206 (67.5%) | 206 (15.3%) |
| **Prior vaccination** | 0 (0.0%) | 232 (77.1%) | 232 (17.2%) |
| **Vaccine doses\*** |  |  |  |
| 1-2 doses | — | 173 (74.6%) | 173 (74.6%) |
| 3 doses | — | 59 (25.4%) | 59 (25.4%) |
| **Time since last vaccine dose\*** |  |  |  |
| <6 months | — | 180 (77.6%) | 180 (77.6%) |
| ≥6 months | — | 52 (22.4%) | 52 (22.4%) |

| Type of vaccines received* |  |  |  |
| --- | --- | --- | --- |
| mRNA | — | 231 (99.6%) | 231 (99.6%) |
| Adenovirus vector | — | 1 (0.4%) | 1 (0.4%) |
| Prior SARS-CoV-2 infection | 0 (0.0%) | 36 (11.8%) | 36 (2.7%) |

Overall, 25.3% (95% CI 22.7–28.0%, n=264) of individuals with Wildtype SARS-CoV-2, 17.2% (11.0– 25.8%, n=17) of Delta-infected, and 13.1% (9.2–18.4%, n=27) of Omicron-infected individuals were affected by PCC six months after their last infection. The proportion of participants with PCC among non-vaccinated individuals infected with the Delta (21.6%, 11.4–37.2%, n=8) and Omicron (21.9%, 11.0– 38.8%, n=7) variants were similar compared to Wildtype infection without prior vaccination. Among individuals that had received at least one vaccination dose prior to infection, 14.8% (8.0–25.7%, n=9) had PCC after Delta, and 11.1% (7.2–16.7%, n=19) after Omicron infection. No clear patterns in PCC-related symptoms were observed across individuals infected with the different variants (Supplementary Figure S2).

When assessing the association between infection with different SARS-CoV-2 variants and prior vaccination with PCC at six months, there was strong evidence for a reduction in the risk among vaccinated individuals infected by the Omicron variant (OR 0.42, 95% CI 0.24–0.68, p=0.0008) compared to non-vaccinated participants infected with Wildtype SARS-CoV-2, based on multivariable logistic regression analyses adjusted for age, sex, presence of comorbidities, hospitalization due to COVID-19, and prior infection (Figure 1). Meanwhile, there was weak evidence for a risk reduction among vaccinated individuals infected with Delta (OR 0.55, 0.25–1.08, p=0.11), but no evidence for a reduction in the risk among non-vaccinated individuals infected by the Delta (OR 0.84, 0.34–1.87, p=0.69) or Omicron (OR 0.87, 0.33–2.06, p=0.77) variant. In sensitivity analyses including variant and vaccination as separate variables, results were similar for the association of Delta (OR 0.92, 0.44–1.83, p=0.83) and Omicron (OR 0.78, 0.35–1.65, p=0.54) infection, as well as prior vaccination with PCC (OR 0.55, 95% CI 0.26–1.19, p=0.12; Supplementary Figure S3).

**Figure 1:**
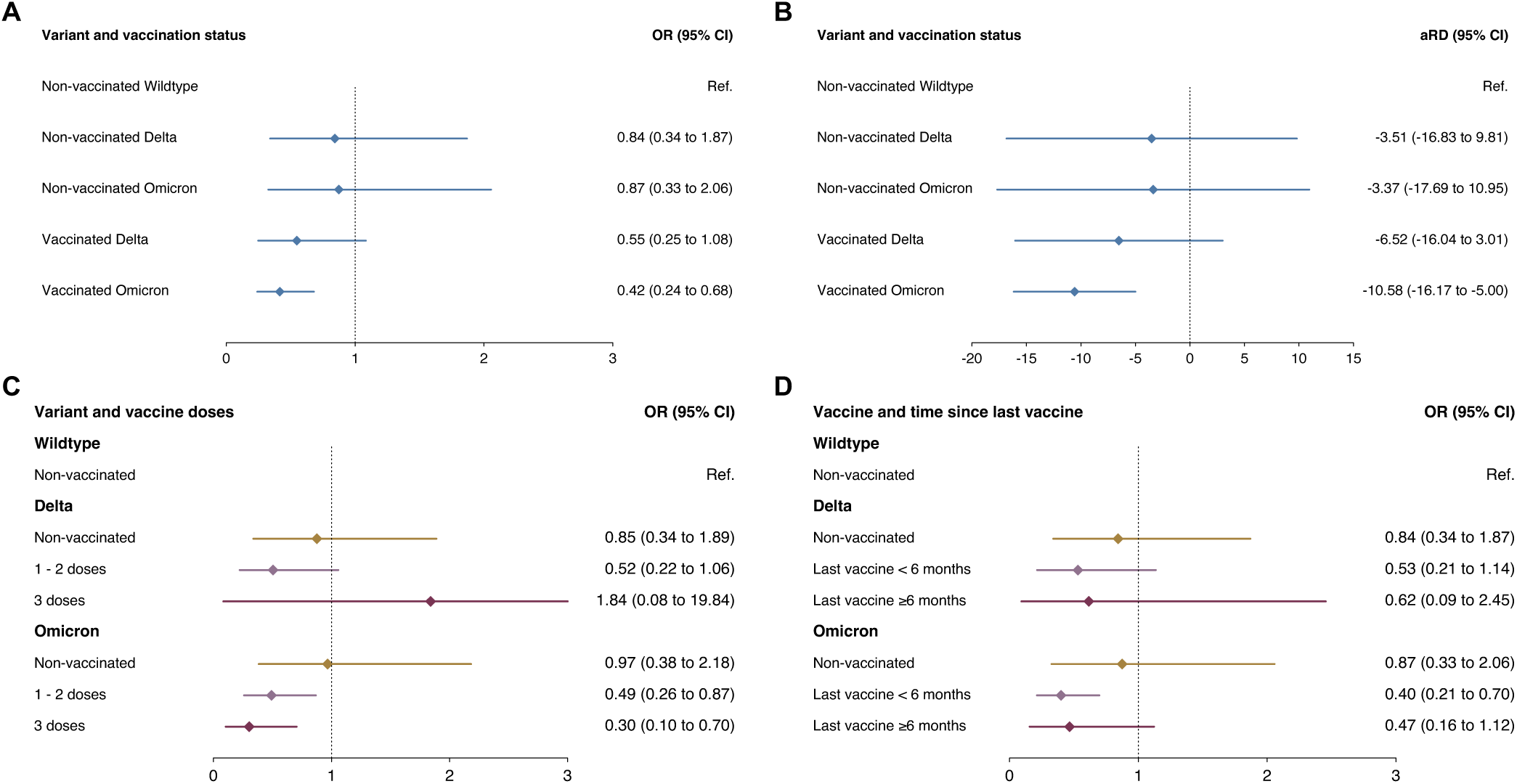
Association of Delta and Omicron SARS-CoV-2 infection and prior vaccination with post COVID-19 syndrome six months after SARS-CoV-2 infection. Panels **A** and **B** show odds ratios and adjusted risk differences among non-vaccinated and vaccinated individuals infected with the Delta or Omicron variant compared to non-vaccinated individuals infected by SARS-CoV-2, based on multivariable logistic regression models adjusted for age, sex, presence of comorbidities, initial hospitalization due to COVID-19, and prior infection. Panels **C** and **D** show odds ratios for having received one or two vaccine doses or three doses prior to infection and for having been vaccinated less than six months prior or six or more months prior to infection with the Delta or Omicron variants, based on multivariable logistic regression models adjusted for age, sex, presence of comorbidities, initial hospitalization due to COVID-19, and prior infection. aRD = adjusted risk difference, CI = confidence interval, OR = odds ratio, Ref. = reference group.

In analyses stratified by the number of vaccine doses received prior to infection, there was no evidence for a difference between individuals that had received one or two vaccine doses and individuals that had received three doses among individuals infected with the Delta variant, while there was a tendency for a stronger effect with three (OR 0.30, 0.10–0.70, p=0.013) compared to one or two doses (OR 0.49, 0.26– 0.87, p=0.020) with the Omicron variant (Figure 1). The risk independent of variant was equally reduced for both one to two doses and three doses compared to non-vaccinated individuals, however with high uncertainty (Supplementary Figure S3). When evaluating the timing since the last vaccine dose at infection, there was no evidence for a difference between individuals that were vaccinated six or more months prior, and individuals vaccinated less than six months prior to infection among individuals infected with the Delta and the Omicron variant (Figure 1). Findings were similar in a sensitivity analysis of associations independent of variants (Supplementary Figure S3).

In terms of the severity of PCC-related symptoms at six months, there were similar patterns across groups infected by different variants and with and without prior vaccination (Figure 2). In accordance with the overall prevalence of PCC, we observed lower prevalences of individuals reporting one to two, three to five, and six or more PCC-related symptoms among vaccinated individuals infected with the Omicron variant compared to other groups. Sensitivity analyses using different severity classifications resulted in similar findings (Supplementary Figure S4). While the risk among vaccinated Omicron-infected individuals was lower for those reporting three or more symptoms, 4% (n=7) still reported such symptoms after six months. In multivariable multinomial logistic regression analyses, there was strong evidence for a reduction in those reporting one or two symptoms among vaccinated Omicron-infected individuals compared to non-vaccinated Wildtype-infected individuals (OR 0.39, 95% CI 0.21–0.74, p=0.0038), while the statistical evidence for a reduction of those reporting three to five (OR 0.44, 0.16–1.19, p=0.11) and six or more symptoms (OR 0.50, 95% CI 0.11–2.21, p=0.36) was insufficient (Table 2, Supplementary Tables S6–8).

**Table 2:** Results from multinomial logistic regression analyses of the association of SARS-CoV-2 variant and vaccination with severity of post COVID-19 condition in terms of symptom count. CI = confidence interval, OR = odds ratio, Ref. = reference group.

| Characteristic | 1-2 symptoms |  | 3-5 symptoms |  | 6+ symptoms |  |
| --- | --- | --- | --- | --- | --- | --- |
|  | OR (95% CI) | p-value | OR (95% CI) | p-value | OR (95% CI) | p-value |
| **Non-vaccinated Wildtype** | Ref. |  | Ref. |  | Ref. |  |
| **Non-vaccinated Delta** | 0.50 (0.15–1.74) | 0.28 | 0.76 (0.15–3.72) | 0.73 | 3.91 (0.97–15.7) | 0.055 |
| **Non-vaccinated Omicron** | 0.80 (0.26–2.45) | 0.69 | 0.76 (0.14–4.08) | 0.75 | 1.95 (0.23–16.6) | 0.54 |
| **Vaccinated Delta** | 0.46 (0.18–1.18) | 0.11 | 0.49 (0.11–2.07) | 0.33 | 1.36 (0.31–6.02) | 0.69 |
| **Vaccinated Omicron** | 0.39 (0.21–0.74) | 0.0038 | 0.44 (0.16–1.19) | 0.11 | 0.50 (0.11–2.21) | 0.36 |

**Figure 2:**
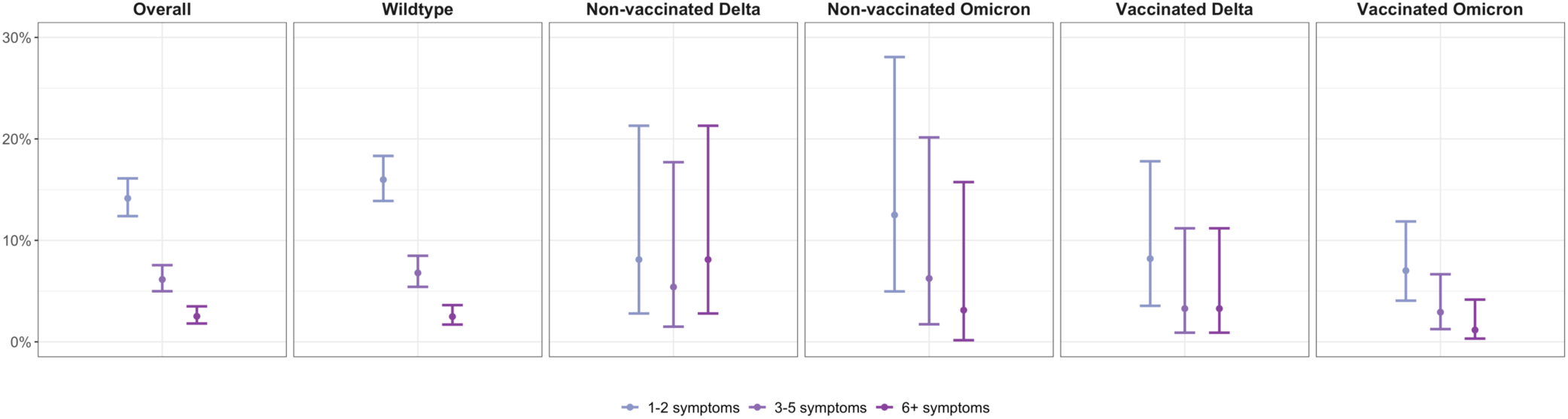
Prevalence of post COVID-19 condition six months after infection at different levels of severity in terms of symptom count. Analyses were stratified by SARS-CoV-2 variant and vaccination status. Points represent point estimate and error bars represent 95% Wilson confidence intervals for estimated proportions.

Cluster analyses resulted in the identification of four clusters with different patterns of PCC-related symptoms (Figure 3). Based on the predominant symptom patterns in each cluster, we categorised them into groups with diverse systemic (n=219 of participants with PCC), neurocognitive (n=47), cardiorespiratory (n=23), and musculoskeletal symptoms (n=19). Meanwhile, certain symptoms were prevalent in all four clusters, such as fatigue, post-exertional malaise, headache, and taste or smell disturbances. In sensitivity analyses assuming five or six clusters, additional groups of individuals predominantly affected by gastrointestinal disturbances or hair loss, and by vertigo or dizziness were identified (Supplementary Figures S5–6). Across infections with different variants, we observed no clear differences in the proportion of participants belonging to each cluster (Figure 3). Female participants were more prevalent in the neurocognitive and cardiorespiratory clusters and older participants and individuals with comorbidities were more prevalent in the cardiorespiratory and musculoskeletal clusters, while the majority of participants with diverse systemic symptoms reported having one PCC-related symptom (Supplementary Table S9).

**Figure 3:**
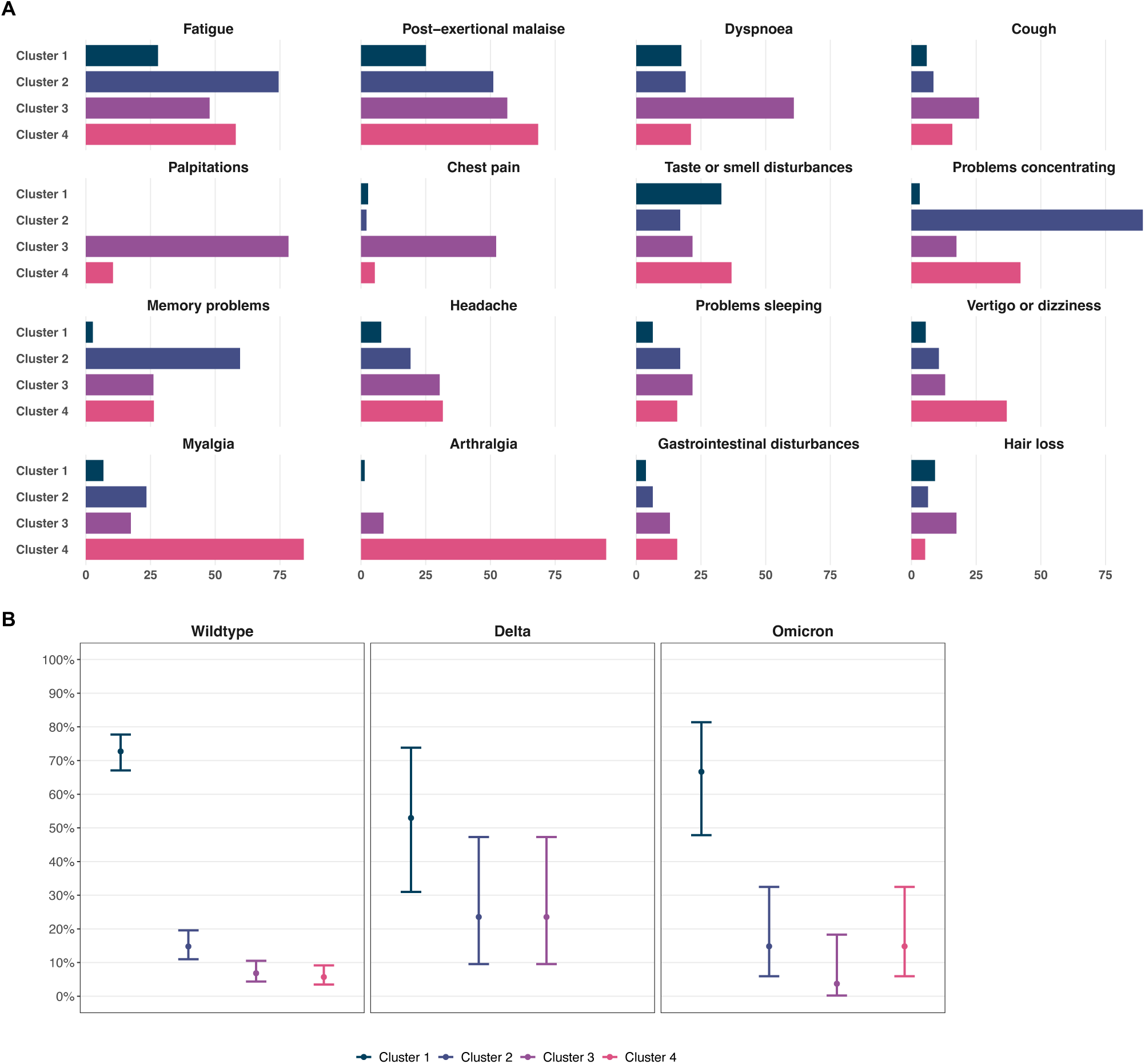
Prevalence of specific post COVID-19 condition-related symptoms six months after SARS-CoV-2 infection across symptom clusters. Four clusters of individuals with post COVID-19 condition at six months after infection were identified based on multiple correspondence and hierarchical cluster analyses, consisting of individuals with (1) diverse systemic symptoms and lower symptom count, and with (2) predominantly neurocognitive, (3) cardiorespiratory, or (4) musculoskeletal symptoms. Panel **A** depicts distributions of specific post COVID-19 condition-related symptoms across clusters. Panel **B** shows the proportion of individuals belonging to each cluster across infections with Wildtype, Delta, and Omicron SARS-CoV-2. Points represent point estimate and error bars represent 95% Wilson confidence intervals for estimated proportions.

In this pooled analysis of 1350 SARS-CoV-2-infected individuals from two population-based cohorts, we found that infection with the Omicron variant and prior vaccination were associated with a lower risk of PCC six months after infection, while the risk among non-vaccinated individuals infected with the Delta or Omicron variant was similar to those infected with Wildtype SARS-CoV-2. We found no differences in the occurrence of PCC between individuals having received one or two vaccine doses and individuals having received three vaccine doses, and between individuals last vaccinated more or less than six months prior infection. Compared to non-vaccinated individuals infected with Wildtype SARS-CoV-2, the severity of PCC symptoms was lower among vaccinated individuals infected with the Omicron variant, but the risk for PCC even of high severity was still present.

To our knowledge, this study is the first to simultaneously evaluate the impact of prior vaccination and infection with the Omicron variant on developing PCC up to six months after infection. In our study, the risk among non-vaccinated individuals infected with the Omicron variant was reduced by approximately 23% compared to those infected by Wildtype SARS-CoV-2, corresponding to an absolute reduction of about 3 in 100 infected individuals. Among vaccinated individuals, the risk was reduced by 45% and 58% for Delta and Omicron infection, respectively, compared to non-vaccinated Wildtype-infected individuals. This corresponds to an absolute risk reduction in Omicron-infected of about 4 in 100 compared to vaccinated Delta-infected individuals and of 10 in 100 compared to non-vaccinated Wildtype-infected individuals. These estimates are in line with a relative risk reduction of 33% between infections with Omicron and prior variants at three months reported in a previous study.^19^ While one further study reported no association of pooled Omicron and Delta infection with PCC compared to Wildtype infection at four weeks,^9^ another reported a stronger risk reduction between Omicron and Delta infection among vaccinated individuals.^18^ They also found a stronger risk reduction among those vaccinated more than three months prior infection compared to those infected less than three months before. In our study, we did not identify a difference depending on whether the last vaccination was received more or less than six months prior to infection. To date, no study investigated potential effects on severity of PCC. Further evidence from representative population-based studies is thus necessary to better estimate the reduction in the risk and severity of PCC in the longer-term while accounting for vaccination. While it cannot be excluded that future variants may bear a higher risk for PCC, this population-based study provides important early evidence on its longer-term risk and severity with the Omicron variant.

There is substantial heterogeneity between existing studies evaluating the effects of vaccination on PCC.^6^ Our findings together with those from others imply that vaccination may reduce the risk of PCC by up to 50%,^7–9,12,13,15–17^ with an estimated relative risk reduction of 52% among Omicron-infected, and 35% among Delta-infected in our study. Despite important differences in study populations, study designs, analytical methods, and definitions of PCC across existing studies, our estimates are in line with previous evidence, albeit of smaller magnitude compared with one other study simultaneously investigating differences between SARS-CoV-2 variants and prior vaccination among non-hospitalised healthcare workers.^9^ The accumulating evidence on potential preventive effects of vaccination on PCC has important implications for vaccination strategies and may be used to inform and positively influence individual decisions regarding booster vaccinations. However, a rigorous evaluation of the evidence, taking into account the substantial heterogeneity in study designs and the time periods during which they took place, is warranted prior to issuing recommendations regarding the prevention of PCC through vaccination.

In cluster analyses, we identified four distinct clusters of PCC-related symptoms across different variants, which we categorised as diverse systemic, neurocognitive, cardiorespiratory, and musculoskeletal symptoms. One study so far has investigated symptom clusters in the context of different SARS-CoV-2 variants.^33^ The authors reported three main emerging PCC symptom clusters (i.e., central neurological, cardiorespiratory, and systemic/inflammatory), while total number of identified clusters and cluster profiles varied across Wildtype, Alpha, and Delta variants. In our study, we found no substantial variation in the prevalence of symptom clusters across infections with different variants, but relevant variation in participant characteristics across clusters. Further studies have investigated the clustering of PCC-related symptoms, leading to the identification of similar groups with neurocognitive, cardiorespiratory, musculoskeletal and pain-related, and systemic or diverse symptoms.^34,35^ Our findings are thus in line with existing evidence, and suggest that some symptoms may be common to all presentations of PCC, while there may be distinct phenotypes of PCC that may also have different pathophysiological explanations or require different clinical management or specific treatment.

This study has several key strengths, including the representative, population-based sample, and the prospective design, alongside a longer-term perspective of PCC six months after infection. Meanwhile, several limitations have to be considered when interpreting the results. First, we pooled data from two different but closely aligned cohorts. Due to the different sampling, recruitment, and timeframes of data collection, there may still be differences between cohorts that we could not fully account for in adjusted analyses. Second, selection may have occurred in both cohorts if participants with long-term symptoms were more likely to be enrolled or complete the questionnaire. However, in the Zurich SARS-CoV-2 Cohort, participants were enrolled prior to the possible occurrence of PCC, and there was minimal loss to follow-up. Corona Immunitas was a highly public, governmentally supported seroprevalence study using a random population sample, from which we included all individuals with diagnosed infection. While some asymptomatically infected individuals in this study may not have sought testing and thus were excluded, we consider the probability of relevant selection bias with respect to the findings to be low. Third, we relied on self-reported measures and could not perform a clinical validation of the relation of symptoms with initial SARS-CoV-2 infection or alternative diagnoses. While information bias cannot be excluded, we consider its potential effect on our findings to be minimal. Fourth, we did not have direct genetic data from viral sequencing and may have misclassified some participants infected with the Delta or Omicron variant based on our cut-off timeframes. Fifth, we determined severity of PCC based on symptom count and EQ-VAS, of which the first may not be a direct correlate of severity and the latter may be influenced by baseline health status. While we tried to account for this in sensitivity analyses leading to broadly similar findings, results may still be confounded. Sixth, vaccinated participants were almost exclusively vaccinated with mRNA-based vaccines, so that our findings may not be fully generalisable to other vaccine types. Seventh, the study was not adequately powered to investigate differences in associations between individuals vaccinated once or twice and individuals that received three vaccine doses, leading to substantial statistical uncertainty. Last, the exploratory cluster analyses bear various limitations inherent in the structure and parametrisation of the model. Identified clusters are probabilistic and sample-bound, and may not coincide with phenotypes of PCC encountered in clinical practice. While our findings align with those by others, further research is necessary to more clearly identify different phenotypes based on their clinical and pathophysiological presentation.

In conclusion, this study demonstrated that infection with Omicron SARS-CoV-2 among individuals with prior vaccination is associated with a substantially reduced risk of PCC at six months post-infection compared to infection with Wildtype SARS-CoV-2 in non-vaccinated individuals, respectively. While the risk of developing PCC appears to persist in the context of vaccination and novel variants, the reduction in risk through vaccination was of greater magnitude than with infection by Delta or Omicron SARS-CoV-2. This information should be considered in vaccination strategies, further vaccine development, and the planning of public health measures for future pandemic waves.

## Data Availability

Deidentified individual participant data underlying the findings of this study will be available for researchers submitting a methodologically sound proposal to achieve the aims of the proposal after peer-reviewed publication of this article.

## Author contributions

TB, DM, JF, and MAP conceived and designed the Zurich SARS-CoV-2 Cohort study. MK, RA, AF, VvW, JF, EA, and MAP conceived and designed the Corona Immunitas seroprevalence study. JF, EA, and MAP supervised the project. TB, DM, MK, RA, AF, and VvW provided administrative, technical, or logistical support for the studies. All authors had full access to the data. DM, TB, and MK accessed and verified the underlying data. TB and DM conducted the statistical analyses. All authors contributed to the interpretation of the findings. TB and DM drafted the first version of the manuscript. All authors critically revised the manuscript for important intellectual content. All authors accept full responsibility for the content of the paper and have seen and approved the final version.

## Data sharing

Deidentified individual participant data underlying the findings of this study will be available for researchers submitting a methodologically sound proposal to achieve the aims of the proposal after publication of this article. Proposals should be directed at the corresponding author (Prof. Dr. Milo A. Puhan).

## Acknowledgements

This study is part of the Corona Immunitas research network, coordinated by the Swiss School of Public Health (SSPH+), and funded by fundraising of SSPH+ including funds of the Swiss Federal Office of Public Health and private funders (ethical guidelines for funding stated by SSPH+ were respected), by funds of the cantons of Switzerland (Vaud, Zurich, and Basel), and by institutional funds of the Universities. Additional funding specific for the two cohorts included in this study was received from the Department of Health of the canton of Zurich, the University of Zurich (UZH) Foundation, and the Swiss Federal Office of Public Health. TB received funding from the European Union’s Horizon 2020 research and innovation programme under the Marie Skłodowska-Curie grant agreement No 801076, through the SSPH+ Global PhD Fellowship Programme in Public Health Sciences (GlobalP3HS) of the SSPH+. DM received funding by the UZH Postdoc Grant, grant no. FK-22-053.

The authors thank the study administration teams in Zurich and Ticino for their dedicated support of the study. Furthermore, the authors thank Hélène E. Aschmann and Anja Domenghino for their contribution to recruitment in the Zurich SARS-CoV-2 Cohort, and Sarah R. Haile for her statistical advice. Last, the authors thank the study participants for their valuable contribution to this project.

