## Supplementary Material for "Post COVID-19 condition after Wildtype, Delta, and Omicron variant SARS-CoV-2 infection and vaccination: pooled analysis of two population-based cohorts"

**Supplementary Figure S1: Flowchart of the enrolment, data collection, and inclusion of participants from the Zurich SARS-CoV-2 Cohort and from Phase 5 of Corona Immunitas in this study.**

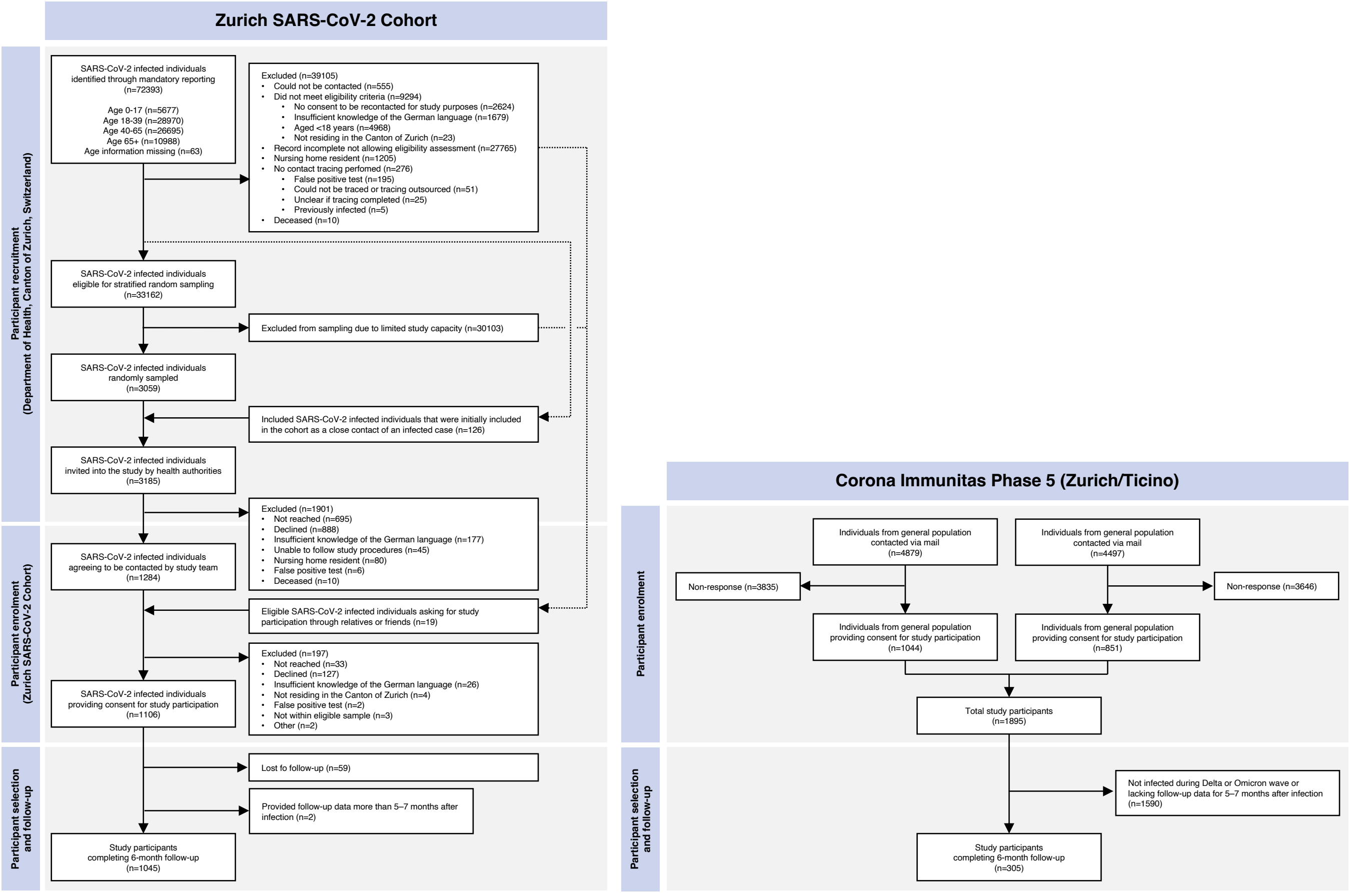

**Supplementary Figure S2: Specific symptoms related to post COVID-19 condition across individuals infected with Wildtype, Delta, and Omicron SARS-CoV-2.** Points represent point estimate and error bars represent 95% Wilson confidence intervals for estimated proportions. GI = gastrointestinal, Prob. = problems, diff. = difficulties.

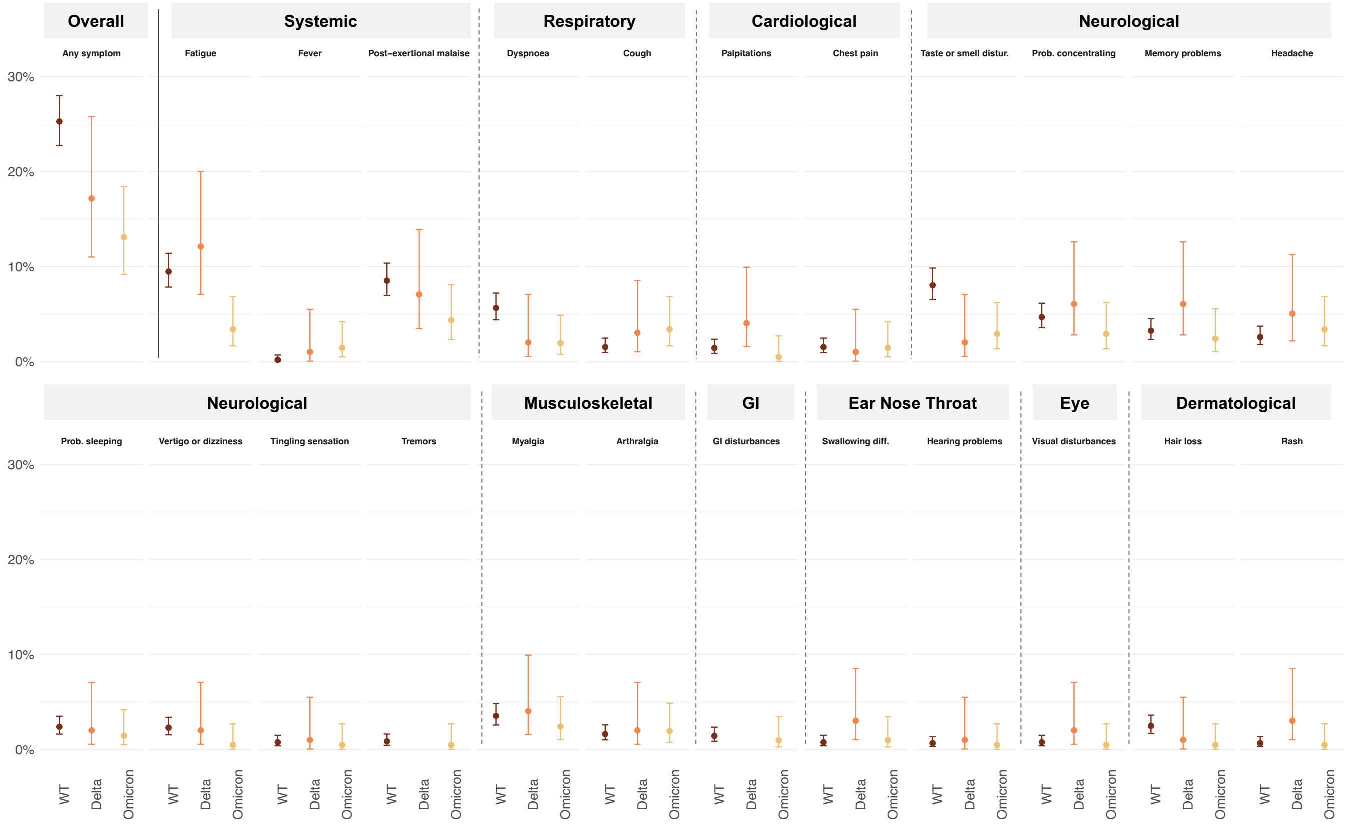

**Supplementary Figure S3: Results from sensitivity analysis of the association of Delta and Omicron SARS-CoV-2 infection, prior vaccination, and prior infection with post COVID-19 syndrome six months after SARS-CoV-2 infection, and of the association of vaccination with post COVID-19 syndrome stratified by number of received vaccine doses and timing of vaccination.** Panel **A** demonstrates independent associations of Delta and Omicron SARS-CoV-2 infection, prior vaccination, and prior infection with post COVID-19 syndrome six months after SARS-CoV-2 infection, based on a multivariable logistic regression model adjusted for age, sex, presence of comorbidities, initial hospitalization due to COVID-19, and prior infection. Panel **B** and **C** show associations of having received one or two vaccine doses or three doses and of having been vaccinated less than six months prior or six or more months prior to infection, based on multivariable logistic regression model adjusted for age, sex, presence of comorbidities, initial hospitalization due to COVID-19, prior infection, and SARS-CoV-2 variant. CI = confidence interval, OR = odds ratio, Ref. = reference group.

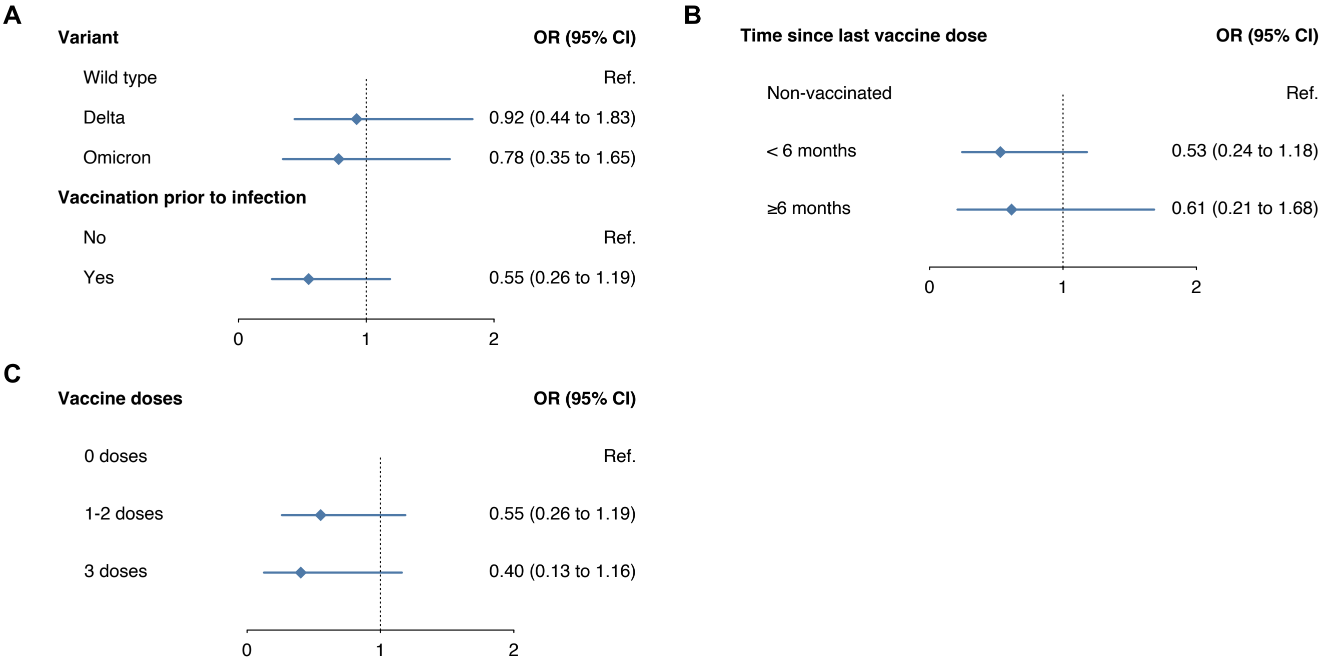

**Supplementary Figure S4: Results from sensitivity analyses regarding the prevalence of post COVID-19 condition six months after infection at different levels of severity.** Panel **A** presents results using symptom count restricted to six symptoms previously found to be in excess among those with post COVID-19 condition compared to the general population (based on Ballouz et al., 2022, https://doi.org/10.1101/2022.06.22.22276746) as severity categories. Panel **B** shows results using current health status based on EQ-VAS scores as severity categories. Panel **C** demonstrates results using current health status based on EQ-VAS scores as severity categories and restricting the analysis to individuals with no reported comorbidities at baseline to account for potential confounding by impaired baseline health status. Points represent point estimate and error bars represent 95% Wilson confidence intervals for estimated proportions.

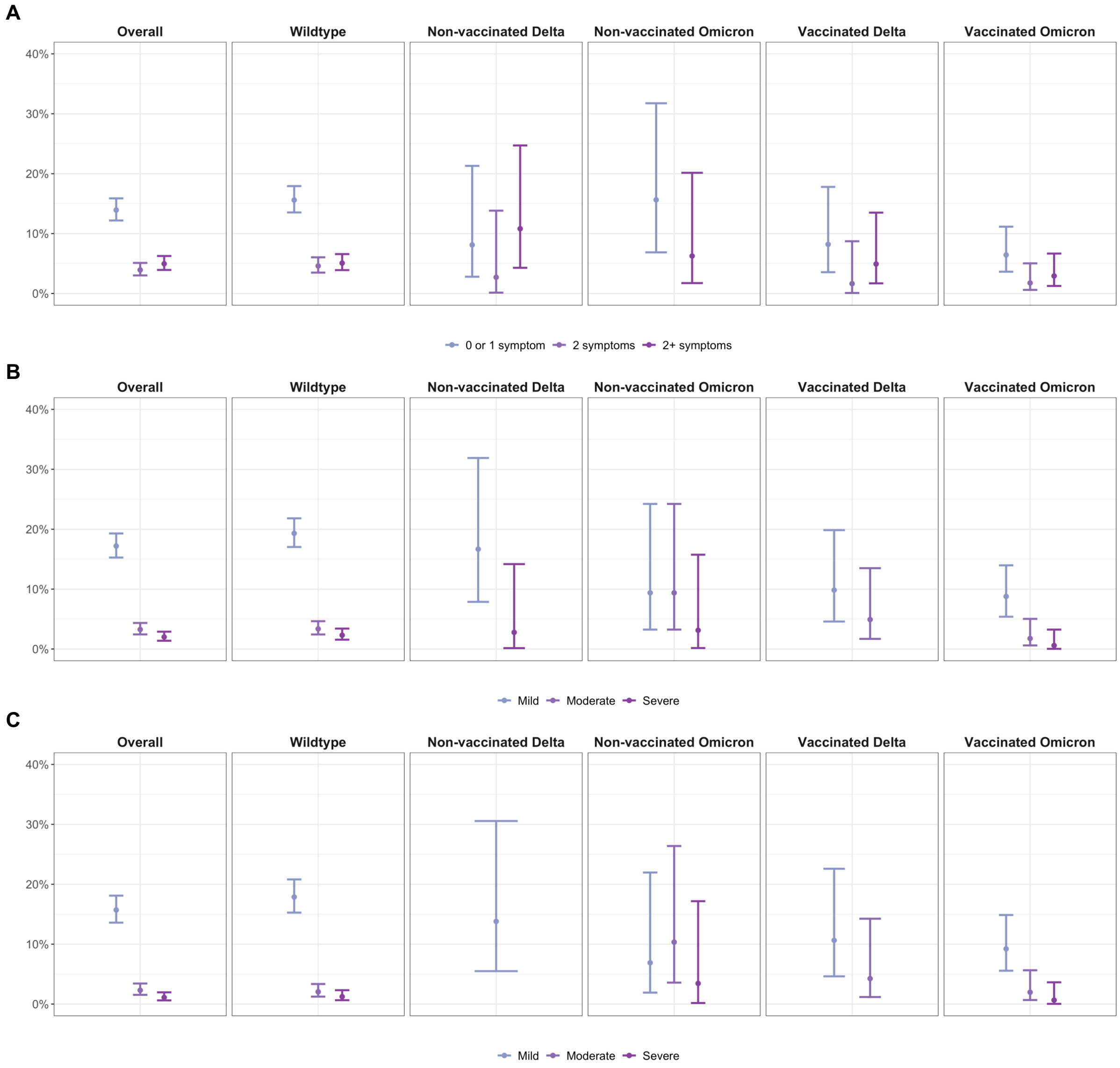

**Supplementary Figure S5: Prevalence of specific post COVID-19 condition-related symptoms six months after SARS-CoV-2 infection across symptom clusters, based on a sensitivity analysis assuming five clusters.** Five clusters of individuals with post COVID-19 condition at six months after infection were identified based on multiple correspondence and hierarchical cluster analyses, consisting of individuals with (1) diverse systemic symptoms and lower symptom count, and with (2) predominantly gastrointestinal symptoms or hair loss, (3) neurocognitive, (4) cardiorespiratory, or (5) musculoskeletal symptoms. Panel **A** depicts distributions of specific post COVID-19 condition-related symptoms across clusters. Panel **B** shows the proportion of individuals belonging to each cluster across infections with Wildtype, Delta, and Omicron SARS-CoV-2. Points represent point estimate and error bars represent 95% Wilson confidence intervals for estimated proportions.

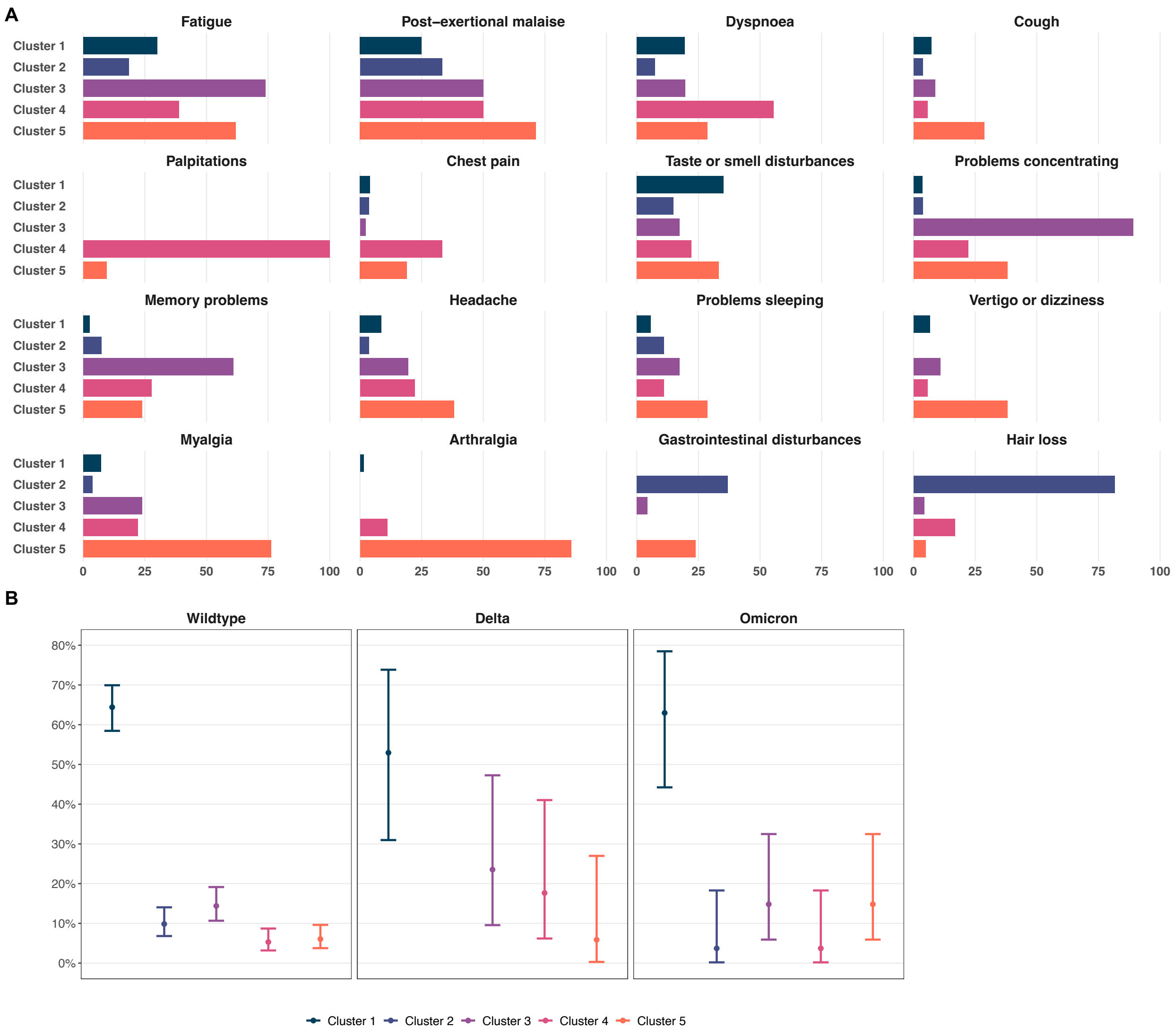

**Supplementary Figure S6: Prevalence of specific post COVID-19 condition-related symptoms six months after SARS-CoV-2 infection across symptom clusters, based on a sensitivity analysis assuming five clusters.** Five clusters of individuals with post COVID-19 condition at six months after infection were identified based on multiple correspondence and hierarchical cluster analyses, consisting of individuals with (1) diverse systemic symptoms and lower symptom count, and with (2) predominantly gastrointestinal disturbances or hair loss, (3) neurocognitive, (4) vertigo or dizziness, (5) cardiorespiratory, or (6) musculoskeletal symptoms. Panel **A** depicts distributions of specific post COVID-19 condition-related symptoms across clusters. Panel **B** shows the proportion of individuals belonging to each cluster across infections with Wildtype, Delta, and Omicron SARS-CoV-2. Points represent point estimate and error bars represent 95% Wilson confidence intervals for estimated proportions.

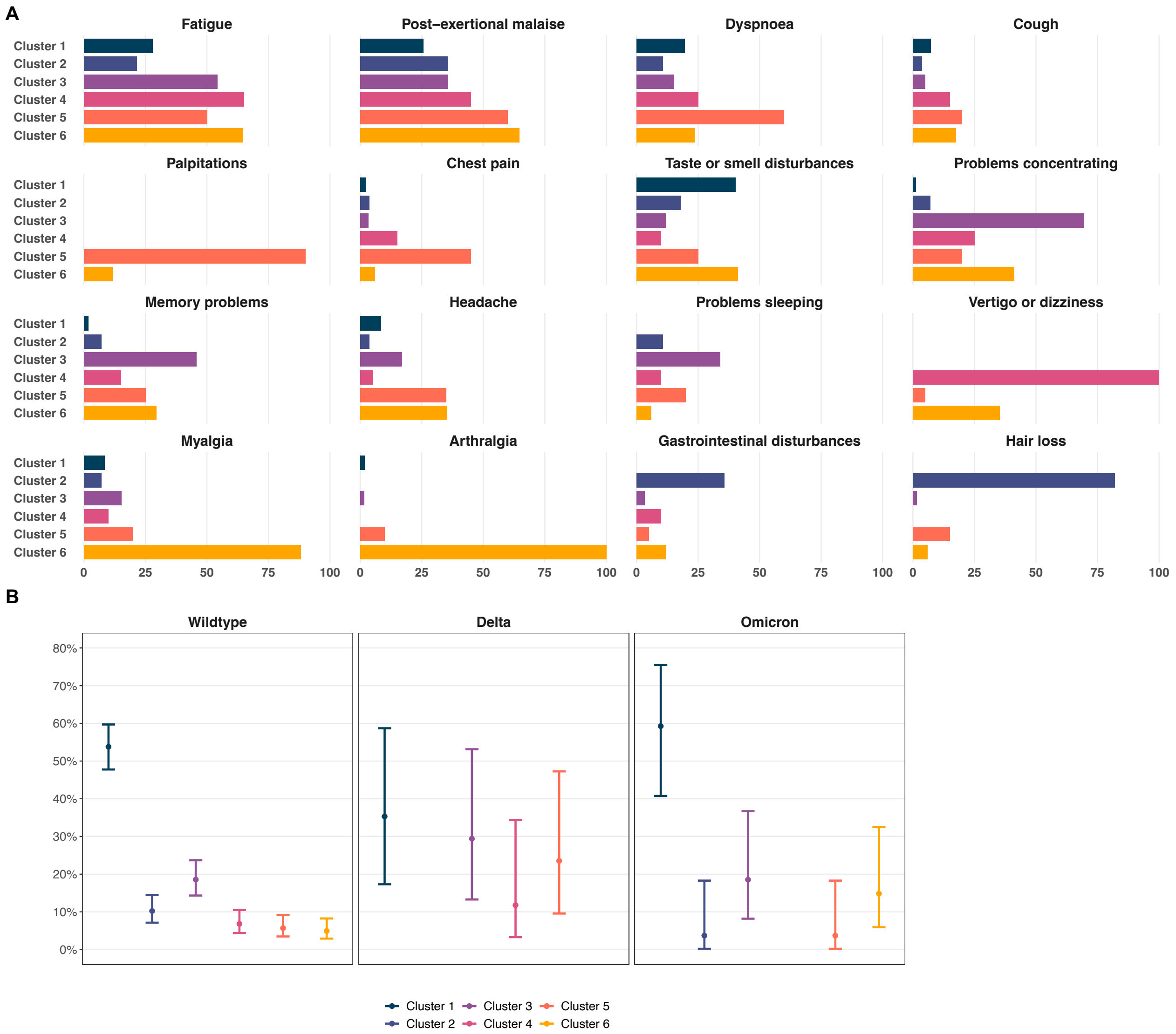

**Supplementary Table S1: Search strategy used in the literature review for the summary of evidence before this study (date of search: Aug 29, 2022).**

| Database | Search strategy | | Number of records |
|---|---|---|---|
| **Medline (OVID)** | 1 | (sars-cov-2 or covid-19 or covid 19 or coronavirus).ti,ab. | 285734 |
| | 2 | COVID-19/ | 182430 |
| | 3 | 1 or 2 | 295332 |
| | 4 | (variant or wildtype or delta or omicron).ti,ab. | 377281 |
| | 5 | (vaccine or vaccination).ti,ab. | 307719 |
| | 6 | 4 or 5 | 677804 |
| | 7 | (post covid-19 condition or post-covid-19 condition or post-acute sequelae or pasc or post covid-19 syndrome or long covid).ti,ab. | 1971 |
| | 8 | 3 and 6 and 7 | 136 |
| | 9 | limit 8 to yr="2020 -Current" | 136 |
| | 10 | animals/ not humans/ | 5006681 |
| | 11 | 9 not 10 | 136 |
| **EMBASE** | #1 | 'sars cov 2':ti,ab OR 'covid 19':ti,ab OR coronavirus:ti,ab | 307106 |
| | #2 | 'coronavirus disease 2019' | 254988 |
| | #3 | #1 OR #2 | 332452 |
| | #4 | variant:ti,ab OR wildtype:ti,ab OR delta:ti,ab OR omicron:ti,ab | 692055 |
| | #5 | vaccine:ti,ab OR vaccination:ti,ab | 369713 |
| | #6 | #4 OR #5 | 1047763 |
| | #7 | 'post covid-19 condition':ti,ab OR 'post-acute sequelae':ti,ab OR pasc:ti,ab OR 'post covid-19 syndrome':ti,ab OR 'long covid':ti,ab | 2400 |
| | #8 | #3 AND #6 AND #7 | 182 |
| | #9 | #8 AND [2020-2022]/py | 182 |
| | #10 | 'animal' NOT 'human' | 4864824 |
| | #11 | #9 NOT #10 | 181 |
| **medRxiv (R code)** | | library(medrxivr)<br>preprint_data <- mx_snapshot()<br>topic1 <- c("COVID-19", "SARS-CoV-2")<br>topic2 <- c("Long Covid", "Post Covid-19 Syndrome", "Post Covid-19 Condition", "post-acute")<br>topic3 <- c("vaccination", "vaccine", "variant", "Omicron", "Delta") | 20 |

| | | | |
|---|---|---|---|
| | | myquery <- list(topic1, topic2, topic3)<br><br>result_list <- mx_search(data = preprint_data, query = myquery)<br><br>mx_export(result_list) | |
| **Result** | | after deduplication | 221 |

**Supplementary Table S2: Eligibility criteria and recruitment timeframes of the Zurich SARS-CoV-2 Cohort and the Corona Immunitas seroprevalence study.**

| | **Zurich SARS-CoV-2 Cohort**<br>**(prospectively recruited participants)*** | **Corona Immunitas**<br>**Zurich & Ticino**<br>**Phase 5** |
|---|---|---|
| **Enrolment timeframe** | Aug 6, 2020 – Jan 26, 2021 | Mar 1, 2022 – Mar 31, 2022 |
| **Sample** | Prospective age-stratified (18–64 years, 65+ years) random sample of all individuals with a diagnosed SARS-CoV-2 infection in the canton of Zurich, Switzerland, based on mandatory laboratory reporting to the cantonal public health authorities (Department of Health of the canton of Zurich), contacted via phone and email. | Representative age-stratified (16–29 years, 30–44 years, 45–64 years, 65+ years) random population sample of the population of the cantons of Zurich and Ticino obtained from the Swiss Federal Statistical Office. |
| **Eligibility criteria** | • Laboratory-confirmed SARS-CoV-2 infection between Aug 6, 2020 and Jan 19, 2021<br>• Residing in the canton of Zurich, Switzerland<br>• Aged 18 years or more<br>• Sufficient knowledge of the German language<br>• Able to follow study procedures<br>• Providing electronic informed consent | • Part of random population sample, responding to study invitation<br>• Resident of the cantons of Zurich or Ticino<br>• Aged 16 years or more<br>• No acute infection with SARS-CoV-2<br>• Providing written informed consent |
| **Sample size** | N=1106<br>The sample size was determined based on the objective of evaluating longer-term health status, symptoms, and medical complications or sequelae of SARS-CoV-2 infection across the full severity spectrum of COVID-19. Based on the knowledge available at the time of designing the study, a sample size of 1200 prospectively recruited individuals was deemed sufficient | N=1894<br>The sample size was determined based on the objectives of the Corona Immunitas seroprevalence study, aiming to evaluate seroprevalence in Switzerland across social and cultural contexts. Based on the sensitivity and specificity of the used antibody test (98% and 99%, respectively) and an expected seroprevalence of ≥90%, a sample size of 200 individuals per age |

| | | |
|---|---|---|
| | to evaluate differences between different groups of interest (e.g., age, disease severity, and sociodemographic differences) and identify rare sequelae after infection. | stratum and canton was determined as sufficient to identify meaningful differences. |

**Legend**: * The Zurich SARS-CoV-2 Cohort consists of two samples of infected individuals. Prospectively recruited participants were enrolled prospectively based on newly reported cases with diagnosed SARS-CoV-2 infection from Aug 6, 2020 to Jan 19, 2021. The study also includes a retrospectively recruited sample for which all individuals diagnosed with SARS-CoV-2 infection between the start of the pandemic in Switzerland (Feb 27, 2020) and start of recruitment (Aug 5, 2020) were contacted. Since testing during the first pandemic wave was restricted in Switzerland and considerations about comparability of populations are warranted, we did not include this sample for the current study. In addition, the cohort includes a case-ascertained sample of close contacts of prospectively recruited individuals, which is also not subject of this study.

**Supplementary Table S3: List of the 23 post COVID-19 condition-related symptoms elicited in the Zurich SARS-CoV-2 Cohort and the Corona Immunitas seroprevalence study questionnaires.**

| **Symptoms**<br>**(translated)** | **Symptoms**<br>**(original wording in German)** |
|---|---|
| In the last 7 days, did you have one or several of the following symptoms, which are not related to a chronic condition or an allergy? (Please select all symptoms that you experienced)<br>• Severe tiredness or exhaustion (fatigue)<br>• Problems breathing or shortness of breath (dyspnoea)<br>• Reduced physical performance (post-exertional malaise)<br>• Cough<br>• Palpitations<br>• Chest pain<br>• Headache<br>• Muscle or bone pain (myalgia)<br>• Joint pain (arthralgia)<br>• Fever<br>• Diarrhea or stool irregularities (gastrointestinal disturbances)<br>• Taste or smell disturbances<br>• Globus sensation or problems swallowing<br>• Tingling or numbness<br>• Trembling<br>• Problems hearing<br>• Vertigo or balance problems<br>• Problems seeing (e.g., flickering, flashes)<br>• Rash<br>• Hair loss<br>• Memory problems<br>• Problems concentrating<br>• Problems sleeping | Hatten Sie in den letzten 7 Tagen eines oder mehrere der folgenden Symptome, welche in keinem Zusammenhang mit einer chronischen Erkrankung oder Allergie stehen? (Bitte geben Sie alle Symptome an, die Sie hatten)<br>• Starke Müdigkeit oder Erschöpfung (Fatigue)<br>• Atembeschwerden oder Kurzatmigkeit<br>• Reduzierte körperliche Leistungsfähigkeit<br>• Husten<br>• Herzklopfen oder Herzstolpern<br>• Brustschmerzen<br>• Kopfschmerzen<br>• Muskel- oder Gliederschmerzen<br>• Gelenkschmerzen<br>• Fieber<br>• Durchfall oder Stuhlunregelmässigkeiten<br>• Störungen des Geruch- oder Geschmacksinns<br>• Einen Kloss im Hals oder Schwierigkeiten zu schlucken<br>• Kribbeln oder Taubheitsgefühl<br>• Zittern<br>• Hörprobleme<br>• Schwindel oder Balancestörungen<br>• Sehstörungen (z.B. Flimmern, Blitze)<br>• Ausschlag<br>• Haarausfall<br>• Gedächtnisprobleme<br>• Konzentrationsprobleme<br>• Schlafprobleme |
| For each of the symptoms present: | Für jedes vorhandene Symptom: |

| | |
|---|---|
| Do you think that this symptom is related to the coronavirus disease "COVID-19" or a complication of the disease? | Denken Sie, dass dieses Symptom mit der Coronavirus-Krankheit "COVID-19" zusammenhängt oder eine Komplikation dieser Krankheit ist? |

**Supplementary Table S4: Detailed participant characteristics of Wildtype, Delta, or Omicron SARS-CoV-2-infected individuals from the Zurich SARS-CoV-2 Cohort and the Corona Immunitas seroprevalence studies (wave V), stratified by study site.** BMI = body mass index, CHF = Swiss Francs, IQR = interquartile range. * Percentages among those that have received at least one vaccine dose prior to infection.

| | **Zurich SARS-CoV-2 Cohort** | **Corona Immunitas** | | **Overall** |
|---|---|---|---|---|
| | | **Ticino** | **Zurich** | |
| | **(N=1045)** | **(N=148)** | **(N=157)** | **(N=1350)** |
| **Timeframe of diagnosis** | Aug 5, 2020 – Jan 19, 2021 | Jul 15, 2021 – Feb 24, 2022 | Jul 26, 2021 – Feb 25, 2022 | Aug 5, 2020 – Feb 25, 2022 |
| **Median follow-up (IQR; days)** | 183.5 (182–186) | 187 (165–196) | 179 (162–193) | 183 (182–186) |
| **Age, median (IQR)** | 51 (35–66) | 41 (30–54) | 43 (31–54) | 48 (34–63) |
| **Age group** | | | | |
| 16-29 | 158 (15.1%) | 37 (25.0%) | 35 (22.3%) | 230 (17.0%) |
| 30-44 | 243 (23.3%) | 50 (33.8%) | 49 (31.2%) | 342 (25.3%) |
| 45-64 | 346 (33.1%) | 49 (33.1%) | 54 (34.4%) | 449 (33.3%) |
| 65+ | 298 (28.5%) | 12 (8.1%) | 19 (12.1%) | 329 (24.4%) |
| **Female sex** | 530 (50.7%) | 96 (64.9%) | 83 (52.9%) | 709 (52.5%) |
| **Presence of chronic comorbidity** | 308 (29.5%) | 23 (15.5%) | 20 (12.7%) | 351 (26.0%) |
| **Smoking status** | | | | |
| Non-smoker | 625 (60.1%) | 98 (66.2%) | 103 (65.6%) | 826 (61.4%) |
| Ex-smoker | 282 (27.1%) | 31 (20.9%) | 31 (19.7%) | 344 (25.6%) |

| | | | | |
|---|---|---|---|---|
| Smoker | 133 (12.8%) | 19 (12.8%) | 23 (14.6%) | 175 (13.0%) |
| *Missing* | *5* | *0* | *0* | *5* |
| **BMI, median (IQR; kg/m2)** | 24.2 (21.9–26.6) | 23.2 (21.3–25.4) | 23.5 (21.6–26.3) | 24.0 (21.7–26.5) |
| **Highest education** | | | | |
| None or mandatory school | 41 (3.9%) | 11 (7.5%) | 7 (4.5%) | 59 (4.4%) |
| Vocational training or specialised baccalaureate | 438 (42.2%) | 84 (57.1%) | 70 (44.9%) | 592 (44.1%) |
| Higher technical school or college | 276 (26.6%) | 9 (6.1%) | 27 (17.3%) | 312 (23.2%) |
| University | 284 (27.3%) | 43 (29.3%) | 52 (33.3%) | 379 (28.2%) |
| *Missing* | *6* | *1* | *1* | *8* |
| **Employment status** | | | | |
| Employed | 668 (64.2%) | 28 (18.9%) | 29 (18.6%) | 725 (53.9%) |
| Retired | 256 (24.6%) | 79 (53.4%) | 102 (65.4%) | 437 (32.5%) |
| Student | 50 (4.8%) | 13 (8.8%) | 19 (12.2%) | 82 (6.1%) |
| Unemployed or other | 67 (6.4%) | 28 (18.9%) | 6 (3.8%) | 101 (7.5%) |
| *Missing* | *4* | *0* | *1* | *5* |
| **Monthly household income** | | | | |
| <6'000 CHF | 330 (33.2%) | 51 (37.2%) | 47 (30.9%) | 428 (33.4%) |
| 6'000 - 12'000 CHF | 443 (44.6%) | 61 (44.5%) | 59 (38.8%) | 563 (43.9%) |
| >12'000 CHF | 221 (22.2%) | 25 (18.2%) | 46 (30.3%) | 292 (22.8%) |
| *Missing* | *51* | *11* | *5* | *67* |
| **Hospitalised due to COVID-19** | 44 (4.2%) | 0 (0.0%) | 2 (1.3%) | 46 (3.4%) |

| | | | | |
|---|---|---|---|---|
| **SARS-CoV-2 variant** | | | | |
| Wildtype | 1045 (100%) | 0 (0.0%) | 0 (0.0%) | 1045 (77.4%) |
| Delta | 0 (0.0%) | 42 (28.4%) | 57 (36.3%) | 99 (7.3%) |
| Omicron | 0 (0.0%) | 106 (71.6%) | 100 (63.7%) | 206 (15.3%) |
| **Prior vaccination** | 0 (0.0%) | 112 (76.7%) | 120 (77.4%) | 232 (17.2%) |
| **Vaccine doses*** | | | | |
| 1-2 doses | – | 87 (77.7%) | 86 (71.7%) | 173 (74.6%) |
| 3 doses | – | 25 (22.3%) | 34 (28.3%) | 59 (25.4%) |
| **Time since last vaccine dose*** | | | | |
| <6 months | – | 87 (77.7%) | 93 (77.5%) | 180 (77.6%) |
| ≥6 months | – | 25 (22.3%) | 27 (22.5%) | 52 (22.4%) |
| **Type of vaccines received*** | | | | |
| mRNA | – | 112 (100%) | 119 (99.2%) | 231 (99.6%) |
| Adenovirus vector | – | 0 | 1 (0.8%) | 1 (0.4%) |
| **Prior SARS-CoV-2 infection** | 0 (0.0%) | 16 (10.8%) | 20 (12.7%) | 36 (2.7%) |

**Supplementary Table S5: Comparison of participant characteristics of Corona Immunitas wave V participants in Zurich and Ticino, Switzerland, with non-included individuals (stratified by infection status).** BMI = body mass index, CHF = Swiss Francs, IQR = interquartile range.

| | **Included participants** | **Non-included individuals** | | |
|---|---|---|---|---|
| | **(N=305)** | **Ever infected (N=634)** | **Never infected (N=956)** | **All (N=1590)** |
| **Age, median (IQR)** | 43 (30–54) | 43 (30–59) | 55 (37–69) | 51 (34–66) |
| **Sex** | | | | |
| female | 179 (58.7%) | 364 (57.4%) | 508 (53.2%) | 872 (54.9%) |
| male | 126 (41.3%) | 269 (42.4%) | 445 (46.6%) | 714 (44.9%) |
| other/diverse | 0 (0.0%) | 1 (0.2%) | 2 (0.2%) | 3 (0.2%) |
| *Missing* | *0* | *0* | *1* | *1* |
| **Presence of chronic comorbidity** | 43 (14.1%) | 140 (22.1%) | 288 (30.2%) | 428 (27.0%) |
| *Missing* | *0* | *0* | *3* | *3* |
| **Smoking status** | | | | |
| Non-smoker | 201 (65.9%) | 404 (63.7%) | 563 (59.2%) | 967 (61.0%) |
| Ex-smoker | 62 (20.3%) | 135 (21.3%) | 230 (24.2%) | 365 (23.0%) |
| Smoker | 42 (13.8%) | 95 (15.0%) | 158 (16.6%) | 253 (16.0%) |
| *Missing* | *0* | *0* | *5* | *5* |
| **BMI, median (IQR; kg/m2)** | 23.3 (21.5–26.0) | 23.9 (21.5–26.6) | 24.1 (21.7–27.4) | 24.0 (21.5–27.1) |
| *Missing* | *0* | *1* | *3* | *4* |
| **Highest education** | | | | |

| | | | | |
|---|---|---|---|---|
| None or mandatory school | 18 (5.9%) | 48 (7.6%) | 94 (10.0%) | 142 (9.0%) |
| Vocational training or specialised baccalaureate | 154 (50.8%) | 291 (46.1%) | 458 (48.5%) | 749 (47.6%) |
| Higher technical school or college | 36 (11.9%) | 118 (18.7%) | 157 (16.6%) | 275 (17.5%) |
| University | 95 (31.4%) | 174 (27.6%) | 235 (24.9%) | 409 (26.0%) |
| *Missing* | *2* | *3* | *12* | *15* |
| **Employment status** | | | | |
| Employed | 57 (18.8%) | 161 (25.5%) | 402 (42.5%) | 563 (35.7%) |
| Retired | 181 (59.5%) | 344 (54.5%) | 411 (43.4%) | 755 (47.9%) |
| Student | 32 (10.5%) | 70 (11.1%) | 71 (7.5%) | 141 (8.9%) |
| Unemployed | 34 (11.2%) | 56 (8.9%) | 62 (6.6%) | 118 (7.5%) |
| *Missing* | *1* | *3* | *10* | *13* |
| **Monthly household income** | | | | |
| <6'000 CHF | 98 (33.9%) | 227 (38.2%) | 399 (44.8%) | 626 (42.2%) |
| 6'000 - 12'000 CHF | 120 (41.5%) | 233 (39.2%) | 345 (38.8%) | 578 (38.9%) |
| >12'000 CHF | 71 (24.6%) | 135 (22.7%) | 146 (16.4%) | 281 (18.9%) |
| *Missing* | *16* | *39* | *66* | *105* |
| **Ever vaccinated** | 233 (76.4%) | 571 (90.1%) | 896 (93.7%) | 1467 (92.3%) |

**Supplementary Table S6: Results from sensitivity analyses of the association of SARS-CoV-2 variant and vaccination with severity of post COVID-19 condition based on multinomial logistic regression models, using symptom count restricted to six symptoms previously found to be in excess among those with post COVID-19 condition compared to the general population as severity categories.** Excess symptoms were defined based on Ballouz et al., 2022 (https://doi.org/10.1101/2022.06.22.22276746). CI = confidence interval, n.e. = not estimable, OR = odds ratio, Ref. = reference group.

| **Characteristic** | **0-1 symptoms** | | **2-3 symptoms** | | **4+ symptoms** | |
|---|---|---|---|---|---|---|
| | **OR (95% CI)** | **p-value** | **OR (95% CI)** | **p-value** | **OR (95% CI)** | **p-value** |
| **Non-vaccinated Wildtype** | Ref. | | Ref. | | Ref. | |
| **Non-vaccinated Delta** | 0.51 (0.15–1.77) | 0.29 | 0.56 (0.06–4.97) | 0.60 | 2.35 (0.69–7.97) | 0.17 |
| **Non-vaccinated Omicron** | 1.03 (0.37–2.89) | 0.96 | n.e. | n.e. | 1.41 (0.27–7.43) | 0.68 |
| **Vaccinated Delta** | 0.48 (0.19–1.22) | 0.12 | 0.34 (0.05–2.56) | 0.30 | 1.03 (0.30–3.50) | 0.96 |
| **Vaccinated Omicron** | 0.37 (0.19–0.71) | 0.0027 | 0.38 (0.11–1.35) | 0.14 | 0.65 (0.24–1.74) | 0.39 |

**Supplementary Table S7: Results from sensitivity analyses of the association of SARS-CoV-2 variant and vaccination with severity of post COVID-19 condition based on multinomial logistic regression models, using current health status based on EQ-VAS scores as severity categories.** CI = confidence interval, n.e. = not estimable, OR = odds ratio, Ref. = reference group.

| **Characteristic** | **Mild (EQ-VAS >70)** | | **Moderate (EQ-VAS 51-70)** | | **Severe (EQ-VAS ≤50)** | |
|---|---|---|---|---|---|---|
| | **OR (95% CI)** | **p-value** | **OR (95% CI)** | **p-value** | **OR (95% CI)** | **p-value** |
| **Non-vaccinated Wildtype** | Ref. | | Ref. | | Ref. | |
| **Non-vaccinated Delta** | 0.81 (0.32–2.10) | 0.67 | n.e. | n.e. | 2.46 (0.30–19.9) | 0.40 |
| **Non-vaccinated Omicron** | 0.50 (0.14–1.73) | 0.27 | 2.02 (0.40–10.3) | 0.40 | 3.52 (0.43–28.9) | 0.24 |
| **Vaccinated Delta** | 0.47 (0.20–1.10) | 0.08 | 1.44 (0.41–5.00) | 0.57 | n.e. | n.e. |
| **Vaccinated Omicron** | 0.41 (0.23–0.72) | 0.002 | 0.49 (0.13–1.84) | 0.29 | 0.41 (0.05–3.18) | 0.39 |

**Supplementary Table S8: Results from sensitivity analyses of the association of SARS-CoV-2 variant and vaccination with severity of post COVID-19 condition based on multinomial logistic regression, using current health status based on EQ-VAS scores as severity categories and restricting the analysis to individuals with no reported comorbidities at baseline to account for potential confounding by impaired baseline health status.** CI = confidence interval, n.e. = not estimable, OR = odds ratio, Ref. = reference group.

| **Characteristic** | **Mild (EQ-VAS >70)** | | **Moderate (EQ-VAS 51-70)** | | **Severe (EQ-VAS ≤50)** | |
|---|---|---|---|---|---|---|
| | **OR (95% CI)** | **p-value** | **OR (95% CI)** | **p-value** | **OR (95% CI)** | **p-value** |
| **Non-vaccinated Wildtype** | Ref. | | Ref. | | Ref. | |
| **Non-vaccinated Delta** | 0.68 (0.22–2.05) | 0.49 | n.e. | n.e. | n.e. | n.e. |
| **Non-vaccinated Omicron** | 0.36 (0.08–1.61) | 0.18 | 2.73 (0.53–14.0) | 0.23 | 4.47 (0.52–38.3) | 0.17 |
| **Vaccinated Delta** | 0.55 (0.21–1.42) | 0.22 | 1.98 (0.42–9.37) | 0.39 | n.e. | n.e. |
| **Vaccinated Omicron** | 0.45 (0.25–0.83) | 0.010 | 0.78 (0.20–3.09) | 0.72 | 0.64 (0.08–5.33) | 0.68 |

**Supplementary Table S9: Participant characteristics among infected individuals with post COVID-19 condition in the four clusters identified through cluster analysis.** Identified clusters were composed of (1) diverse systemic, (2) cardiorespiratory, (3) neurocognitive, and (4) musculoskeletal symptoms. BMI = body mass index, IQR = interquartile range.

| | **Cluster 1** | **Cluster 2** | **Cluster 3** | **Cluster 4** |
|---|---|---|---|---|
| | **(N=219)** | **(N=47)** | **(N=23)** | **(N=19)** |
| **Age, median (IQR)** | 52 (37–66) | 49 (34.5–60) | 56 (42–67.5) | 59 (47.5–76.5) |
| **Female sex** | 119 (54.3%) | 33 (70.2%) | 18 (78.3%) | 11 (57.9%) |
| **Presence of chronic comorbidity** | 75 (34.2%) | 15 (31.9%) | 12 (52.2%) | 10 (52.6%) |
| **Smoking status** | | | | |
| Non-smoker | 120 (55.3%) | 27 (57.4%) | 14 (60.9%) | 8 (42.1%) |
| Ex-smoker | 65 (30.0%) | 7 (14.9%) | 6 (26.1%) | 6 (31.6%) |
| Smoker | 32 (14.7%) | 13 (27.7%) | 3 (13.0%) | 5 (26.3%) |
| *Missing* | *2* | *0* | *0* | *0* |
| **Variant** | | | | |
| Wildtype | 192 (87.7%) | 39 (83.0%) | 18 (78.3%) | 15 (78.9%) |
| Delta | 9 (4.1%) | 4 (8.5%) | 4 (17.4%) | 0 (0.0%) |
| Omicron | 18 (8.2%) | 4 (8.5%) | 1 (4.3%) | 4 (21.1%) |
| **Prior vaccination** | 17 (7.8%) | 7 (14.9%) | 1 (4.3%) | 3 (15.8%) |
| **Symptom count, median (IQR)** | 1 (1–2) | 4 (3–6) | 5 (4–7) | 5 (4–9) |
| 1-2 symptoms | 179 (81.7%) | 10 (21.3%) | 2 (8.7%) | 0 (0.0%) |
| 3-5 symptoms | 39 (17.8%) | 22 (46.8%) | 11 (47.8%) | 11 (57.9%) |
| 6+ symptoms | 1 (0.5%) | 15 (31.9%) | 10 (43.5%) | 8 (42.1%) |
